# Improving machine learning and deep learning models for 30-day ICU readmission prediction using Ensemble Bayesian Model Averaging

**DOI:** 10.64898/2026.05.11.26352879

**Authors:** Emanuele Koumantakis, Konstantina Remoundou, Carmen Fava, Ioanna Roussaki, Alessia Visconti, Paola Berchialla

## Abstract

Intensive Care Unit (ICU) readmissions are associated with adverse clinical outcomes and increased healthcare costs. Although existing models for predicting 30-day ICU readmission show high predictive performance, they fail to account for model uncertainty, potentially resulting in overconfident and unreliable decision-making. We propose a novel Ensemble Bayesian Model Averaging (EBMA)-based framework which balances predictive discrimination with uncertainty by penalizing models that are confident but incorrect. It achieved excellent calibration (Brier score = 0.051), while maintaining discriminatory performance comparable to or exceeding that of the best individual models (AUROC > 0.716). These findings suggest that our EBMA-based framework provides a more robust and clinically reliable approach for ICU readmission prediction and decision support.

## Introduction

Unplanned intensive care unit (ICU) readmission is considered a quality indicator, and has been associated with worse outcomes, including higher mortality, longer length of stay, and greater costs, making reliable risk stratification clinically and operationally important^1,2^. Existing predictive models for ICU readmission, ranging from traditional statistical and machine learning to deep learning approaches, have yet to achieve optimal performance in the general ICU population^3–5^. Moreover, even when optimal predictive accuracy is achieved, the clinical utility of these models is often limited by poor calibration (*i*.*e*., low agreement between predicted probabilities and observed frequencies) and overconfident predictions, which can mislead clinicians^6^.

Combining multiple predictive models to improve overall performance through ensemble learning is a well-established and often effective strategy^7,8^. However, the most used ensemble approaches, *e*.*g*., bagging^7^, boosting^9^, and stacking^10^, overlook model uncertainty, still potentially leading to overconfident decision-making^11^. In contrast, Ensemble Bayesian Model Averaging (EBMA)^12–14^ provides a robust framework for incorporating model uncertainty. It calculates the weighted average of predictions from multiple models under the assumption that none of them truly generated the data, but all are potential proxies of the true data-generating model. EBMA rewards discriminative power, yet penalizes models that are confident but incorrect, optimizing ensemble weights so that the output more closely reflects the true probability of the event^13,14^.

Here, we show that combining predictions from models ranging from traditional statistical to machine learning and deep learning approaches within an EBMA-based framework **(Figure 1)** enhances both ICU readmission prediction and model calibration, providing a more robust basis for clinical decision-making.

**Figure 1.**
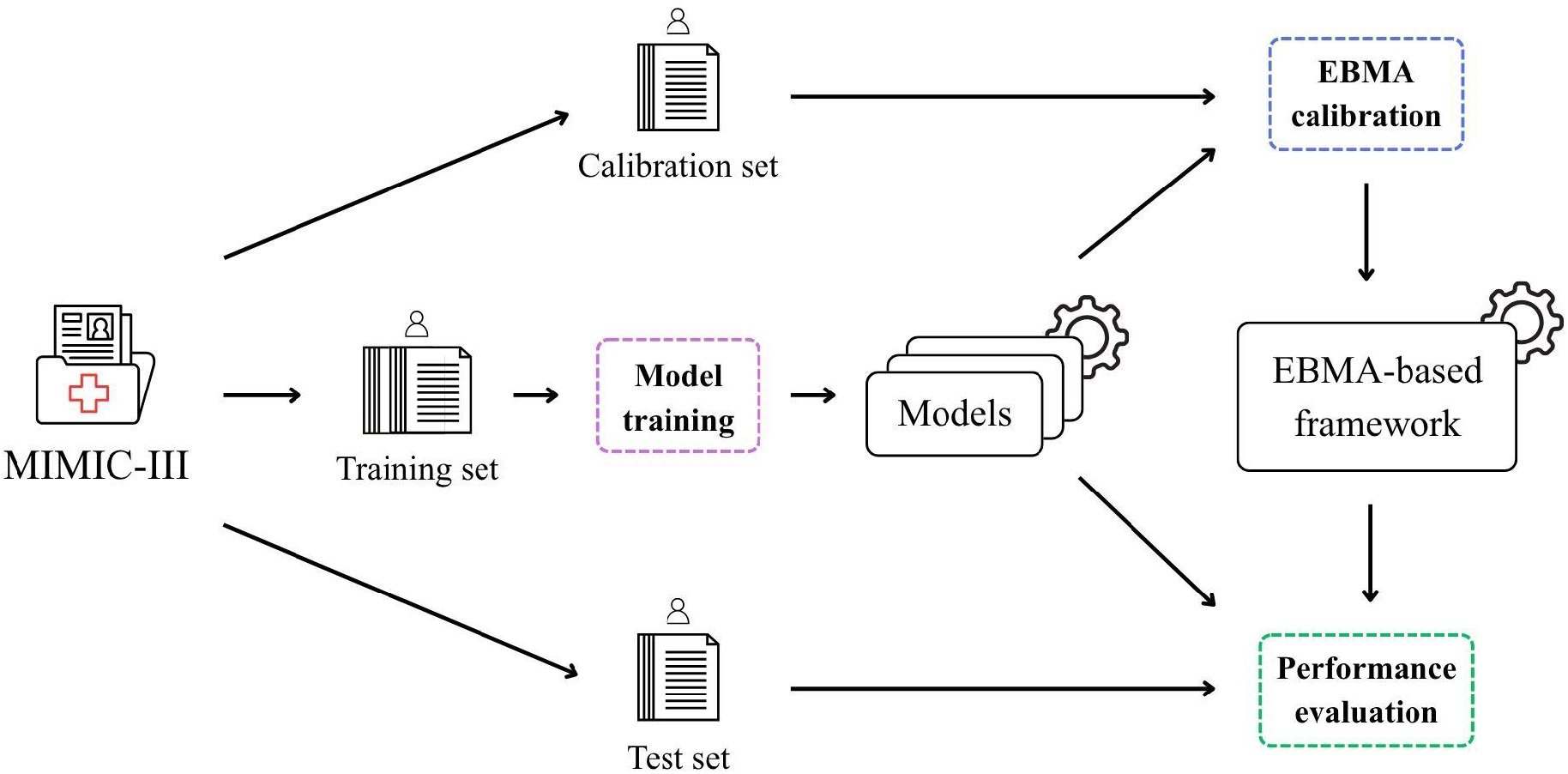
EBMA-based framework overview. The MIMIC-III ICU data for 32,784 patients was split into training (80%), calibration (13%), and test set (7%). The training set was used to train 25 models for predicting 30-day ICU readmission (magenta box). These models were then used to generate prediction probabilities for patients in the calibration set, which were used to calibrate the EBMA weights (blue box), and in the test set, which were used to evaluate the individual model performance (green box). The test set was also used to evaluate the performance of the EBMA-based framework. *Abbreviations: EBMA, Ensemble Bayesian Model Averaging; MIMIC-III, Medical Information Mart for Intensive Care III*.

## Results

### MIMIC-III ICU readmission dataset

After excluding patients who died during the hospital stay or within 30 days after discharge without ICU readmission, our dataset included 32,784 patients, 2,040 (6.2%) of whom would be readmitted to ICU. The mean patient age was 62.6 years old (standard deviation [SD] = 17.5 years old), and 14,001 of them were females (42.7%). We split these data into three subsets for training (n=26,227, 80%), calibration (n=4,371, 13%), and testing (n=2,186, 7%). Each subset maintained the same proportion of ICU readmitted patients as well as a comparable age, with the calibration set including a marginally larger proportion of female patients compared to the training and test set **(Table 1)**. The training set was used exclusively to train the 25 prediction models, while the calibration set was used to quantify model diversity and to calibrate the EBMA-based framework. All models were evaluated on the same test set, which was not used at any stage of model development.

**Table 1.** Sample characteristics. The preprocessed MIMIC-III dataset was divided into 80% training, 13% calibration, and 7% test set. Differences between these sets were calculated using the ANOVA (age) and χ^2^ test (sex, readmission). The statistically significant difference in the proportion of females is likely due to the large sample size, as highlighted by the marginal difference in magnitude.

|  | Overall<br>(N=32784) | Training<br>(N=26,227) | Calibration<br>(N=4,371) | Test<br>(N=2,186) | P |
| --- | --- | --- | --- | --- | --- |
| **Sex** |  |  |  |  |  |
| Female | 14,001<br>(42.7%) | 11,142<br>(42.5%) | 1,957<br>(44.8%) | 902<br>(41.3%) | 0.007 |
| Male | 18,783<br>(57.3%) | 15,085<br>(57.5%) | 2,414<br>(55.2%) | 1,284<br>(58.7%) |  |
| **Age (years)** | 62.6 ± 17.5 | 62.5 ± 17.5 | 62.9 ± 17.3 | 63.3 ± 17.8 | 0.08 |
| **Readmission** |  |  |  |  |  |
| No | 30,744<br>(93.8%) | 24,590<br>(93.8%) | 4,089<br>(93.5%) | 2,065<br>(94.5%) | 0.34 |
| Yes | 2,040<br>(6.2%) | 1,637<br>(6.2%) | 282<br>(6.5%) | 121<br>(5.5%) |  |

### Performances of individual models

Discrimination capacity of the individual approaches, as evaluated by the area under the receiver operating characteristic curve (AUROC), was highly variable, ranging from 0.44 (CNN + LSTM, in Lin at al.^15^) to 0.72 (RNN concatenated Δtime + Attention, in Barbieri et al.^16^; **Supplementary Table 1**). Interestingly, the machine learning and deep learning models showed similar performance (mean AUROC= 0.66 ± 0.03 and 0.67 ± 0.09, respectively), suggesting that simpler and less computationally expensive architectures remain highly competitive. This contrasts with findings from our systematic review^5^ where we observed a median AUROC improvement of 11% (interquartile range [IQR] = 3.8%;19.6%) when using deep learning instead of machine learning approaches. To explain this discrepancy, we explored whether the AUROC obtained using our slightly different training and test set partition and a definition of outcome that did not include mortality was consistent with the values reported in the original studies. Surprisingly, all approaches apart from TeDi-BERT, in Agmon et al.^17^, showed decreased performance (median AUROC difference=-5,1%, IQR=-9.9%;-3.4%; **Supplementary Table 1**). This decrease was particularly important for the approaches presented in Lin et al.^15^, (median AUROC difference=-10.3%, IQR=-28.8%;-9.4%), with the three deep learning approaches showing the largest reduction (**Supplementary Table 1**), likely resulting from an increased class imbalance derived from the exclusion of 30-day mortality from the outcome definition. Notably, despite this variation in outcome definition, TeDi-BERT maintained strong performance.

Similarly, calibration highly differed between models, with a median Brier Score of 0.176 (IQR=0.148;0.232, range: 0.051-0.507; **Supplementary Table 1**), with the majority of the models (n=20, 80%) showing a non-optimal calibration, as represented by a Brier Score over 0.1^18^. Notably, among deep learning models, optimal calibration was obtained only by TeDi-BERT. Both deep learning and machine learning models produced predictions concentrated in the moderate-to-high-probability range (0.20-0.60), substantially exceeding the expected outcome prevalence in the test set (**Supplementary Figure 1**) and confirming a systematic overconfidence^6^.

### Ensemble diversity

Ensemble performance improves with greater prediction diversity^19^. To quantify the degree of diversity available for inclusion in the EBMA-based framework, we generated the predicted probabilities of ICU readmission for the patients in the calibration set and calculated their pairwise Pearson correlation coefficient. We used the calibration set because it was not used during model training, but would actually determine the ensemble weight in the ensemble calibration step. Deep learning models from Barbieri et al. (n=13) showed high positive correlation (mean Pearson’s correlation coefficient ρ = 0.81, SD=0.07; **Supplementary Figure 2**). As expected, models that performed poorly, as those developed by Lin et al^15^, showed only weak correlation not only with other models, but also among themselves. Similarly, traditional scores, which follow different modelling techniques, also showed weak correlation with all other models. Taken together, these observations suggest that ensemble diversity will arise mostly from weaker models and traditional scores.

### Performance of the EBMA-based framework

The EBMA-based framework demonstrated good discriminative performance on the test set, achieving an AUROC of 0.728. Although this was only marginally higher than that of the best individual model (RNN concatenated Δtime + Attention^16^; AUROC = 0.716, improvement = 1.7%), the ensemble achieved excellent calibration, with a Brier score of 0.051 and predicted probabilities close to the observed real frequencies (**Supplementary Figure 3**).

EBMA weights can be interpreted, in a Bayesian sense, as the posterior probability that a given model produces the best prediction for any particular observation^13^. Under this interpretation, the RNN (ODE time decay) + Attention model was the strongest individual contributor to the ensemble (w=0.113, 95% Credible Interval [CrI]: 0.007-0.307), followed by ODE + RNN + Attention (w=0.086, 95% CrI: 0.003-0.293) and RNN (concatenated Δtime) + Attention (w=0.078, 95% CrI: 0.003–0.256, **Figure 2, Supplementary Table 2**). Notably, the highest-weighted models showed wide 95% CrI (**Supplementary Figure 4, Supplementary Table 2**), spanning more than 0.2 weight units. This suggests that, although these architectures were identified as the most informative contributors, considerable uncertainty remained regarding their precise relative contribution.

**Figure 2.**
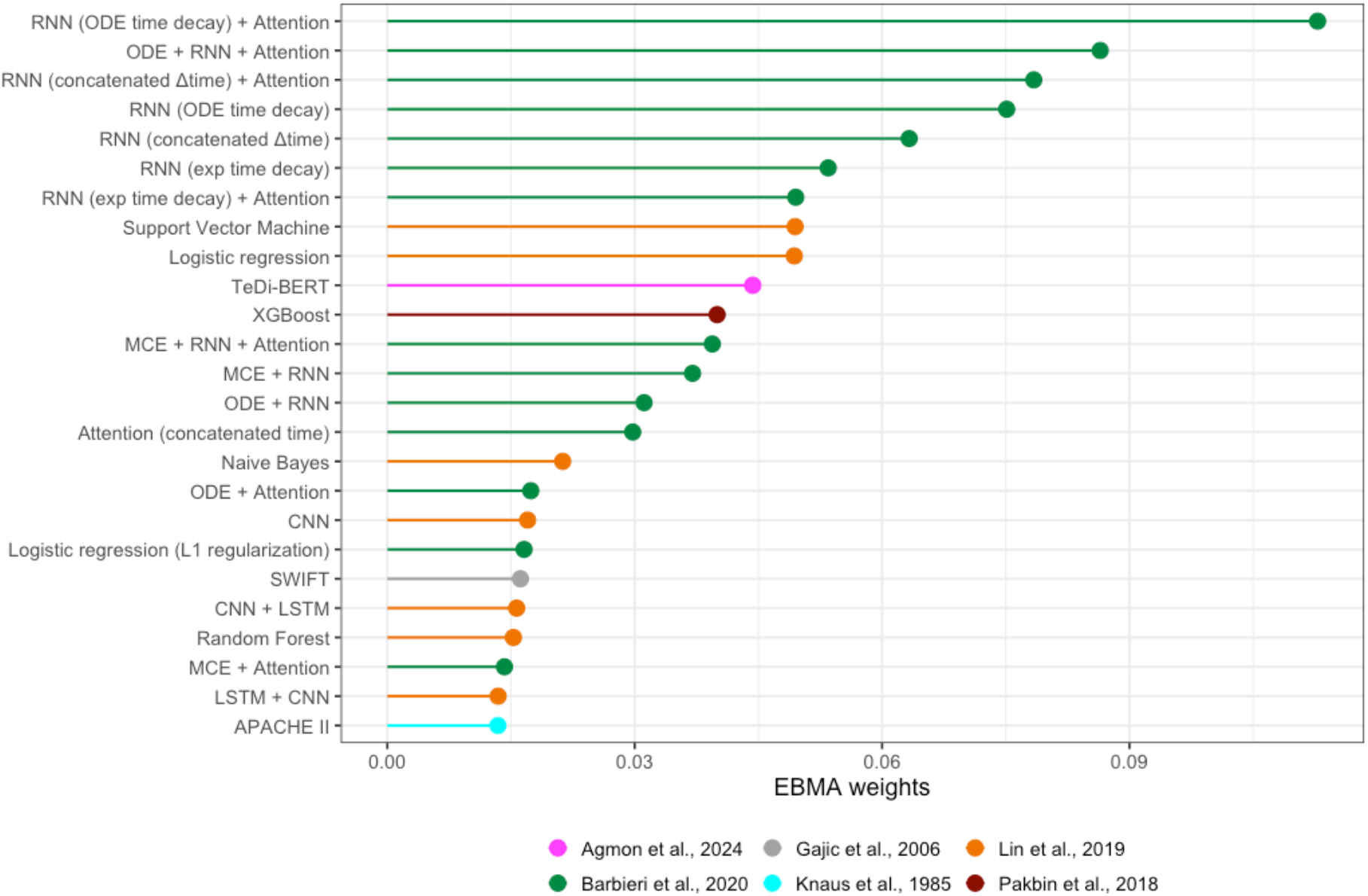
Model predictive performance (weights) in the EBMA-based framework. The lollipop plot shows the weight assigned by the ensemble calibration step to each model. Colours indicate the original studies. *Abbreviations: APACHE II, Acute Physiologic Assessment and Chronic Health Evaluation II; CNN, Convolutional Neural Network; LSTM, Long Short-Term Memory; MCE, Medical Concept Embeddings; ODE, Ordinary Differential Equation; RNN, Recurrent Neural Network; SWIFT, Stability and Workload Index for Transfer; TeDi-BERT, Temporal Distribution matching training method, applied to Bidirectional Encoder Representations from Transformers; XGBoost, eXtreme Gradient Boosting*.

More than half of the models (n=15, 60%) obtained weights that were equal to or lower than the uninformative uniform prior (1/25 = 0.04), contributing minimally to the final prediction. To understand whether removing these models would reduce computational burden without affecting the EBMA-based framework performance, we re-calibrated including only models with weights above 0.04. This retained TeDi-BERT, the logistic regression and the support vector machine from Lin et al.^15^, and seven deep learning models from Barbieri et al.^16^. As expected, performance remained stable achieving an AUROC of 0.731 and a Brier score of 0.051. Interestingly, model diversity remained limited, with a mean Pearson ρ of 0.79 (SD=0.11). We thus investigated whether restricting the ensemble to a smaller set of more diverse models, regardless of their weights, would influence performance. We re-calibrate the EBMA-based framework excluding models whose pairwise Pearson correlation coefficient was larger than 0.8, retaining the models with the highest weight in the original ensemble. The resulting ensemble included 15 models, which offered a good coverage of the different modelling techniques (**Supplementary Table 2**), and yielded similar performance (AUROC=0.727, Brier score=0.051). These results suggest that the ensemble framework can effectively take advantage of the complementary strengths of any available models, including very weak classifiers. Following this observation, we stress-tested this reduced ensemble re-calibrating it after removing the strongest individual contributor (RNN ODE time decay + Attention, w=0.454, 95% CrI=0.277-0.629). We obtained exceptional stability, with an AUROC of 0.716 and a Brier score of 0.051.

## Discussion

It is well recognized that well-calibrated probabilities may be more valuable for clinical decision-making processes than high discrimination alone^20^. Indeed, poor calibration leads to overconfidence and low reliability Here, to address this issue, we implemented a novel EBMA-based framework which achieved good calibration and provided 30-day readmission prediction probabilities that were more reliable than those from the best-performing individual models. This was evidenced by a near-zero Brier Score, achieved without compromising discrimination, as measured by the AUROC. Notably, these results remained robust even in reduced sets of models and, interestingly, even when removing the strongest individual ensemble contributor. Furthermore, our full Bayesian framework estimates the posterior probability of each individual model as the best possible for each observation, providing a rigorous quantification of uncertainty. This helps mitigate overconfidence, which has an important impact, especially for deep learning models^6^.

Additionally, the EBMA-based framework provided a discriminative performance floor, being at minimum equivalent to the best-performing individual model, with our marginal increase in AUROC likely reflecting a lack of diversity among the individual models included in the ensemble.

A key strength of this work is that the candidate models were drawn from previously published studies. We attempted to reproduce both the proposed architectures and the data preprocessing pipelines as faithfully as possible, based on the information and materials made available by the original developers. All analyses were performed on the same dataset on which the models were developed, MIMIC–III, which is high-quality and widely used in critical-care algorithmic research^5^. Additionally, whenever predictions from a new model will be available, integration in our framework is straightforward, and requires little compute and could be run on a standard personal computer.

This study has some limitations. In our recent systematic review, we revealed that despite the proliferation of algorithms to address the ICU readmission prediction task, the overall quality of evidence remains poor^5^. The lack of publicly available code, and, thus, of technical reproducibility, determined a high risk of bias, which limited the number of models that we could include in our ensemble. Therefore, the evaluated set of models represents only a subset of architectures proposed in the ICU readmission literature, and our results may not fully reflect the entire landscape of available approaches. Second, despite efforts to replicate the published methods, some adaptations were necessary to align implementations with our local data partitioning and to enforce a uniform outcome definition across models. Such changes have affected performance of the individual models and, presumably, of our framework.

In summary, by using an EBMA-based framework, we moved away from the “winner-takes-all” mentality of developing yet another standalone model. Instead, we proposed a framework that combines existing predictive knowledge. The proposed ensemble weights models based on their relevance, providing a measure of the uncertainty behind weighting assignment, and mitigating the overconfidence that often leads to performance drop when models are applied to new, unseen patients. Future research should evaluate robustness via external validation and explore strategies to enhance model diversity for greater ensemble gains.

## Methods

### ICU dataset

We used the Medical Information Mart for Intensive Care III (MIMIC-III) database (v1.4), which includes data from 54,423 hospital admissions of patients aged ≥16 years, admitted to the ICUs of the Beth Israel Deaconess Medical Center in Boston, Massachusetts, USA^21^. Patient data span admissions from 2001 to 2012, and the end of follow-up corresponds to the date of hospital discharge or in-hospital death. MIMIC-III integrates detailed clinical information derived from routine care, including demographics, vital signs, laboratory test results, medications, procedures, and clinical notes^21^. We included the first ICU admission for each adult patient aged 18 years or older. Following consensus in the definition of outcome and exclusion criteria^5^, we predicted which patients would be readmitted to the ICU within 30 days from discharge, and excluded patients who died during the hospital stay or within 30 days after discharge without being readmitted to the ICU. We split the resulting dataset into training, calibration, and test set. The training set included the first 80% of the patients, as sorted by their unique patient IDs. The remaining 20% was split randomly between calibration (13%) and test (7%) set.

### Selection of models for the prediction of ICU readmission

#### Deep Learning models

Through a recent systematic review^5^, we retrieved three studies that described a total of 17 deep learning models for ICU readmission prediction, all with publicly available codebases covering data preprocessing and model training^15–17^.

Lin et al.^15^ developed and trained three models: a Convolutional Neural Network (CNN)-based architecture, and two combinations of Long Short-Term Memory (LSTM) and CNN modules. CNNs are optimized for grid-structured data, such as images, but are also suitable for longitudinal electronic health records data. Convolution was performed on the time axis with a 48-hour time window, and the computed feature maps were concatenated to a dense decision layer with one output neuron activated by a sigmoid function. LSTMs can handle the interdependence of events as well as the temporal irregularity and sequential nature of ICU patient data, which evolve throughout an ICU stay. Lin et al.^15^ used a bidirectional LSTM with an additional LSTM layer, followed by a dense decision layer with one output neuron activated by a sigmoid function.

Barbieri et al.^16^, proposed 13 models based on Recurrent Neural Networks (RNNs), modified to learn from irregular, timestamped features. Diagnosis, procedures, medications, and vital sign codes were mapped into embeddings, which were then processed either by recurrent layers, attention mechanisms, or models using neural Ordinary Differential Equations (ODEs).

Agmon et al.^17^ introduced a transformer-based architecture: using the temporal distribution matching training method, applied to Bidirectional Encoder Representations from Transformers (TeDi-BERT). TeDi-BERT was pre-trained on PubMed abstracts of clinical trials between 2010 and 2018 to learn time-aware textual representations, providing representation that are more robust to changes over time in biomedical language and concept usage.

To preserve the original methodologies, we reproduced their preprocessing pipelines and used the same feature sets. However, we adapted the publicly available codebases to ensure that all models were trained and evaluated on the same subsets of patients (*i*.*e*., the training and test set, respectively), noting that for the models in Lin et al.’s^15^, the training set was further randomly divided in a 7:1 ratio, with the smaller portion serving as a validation set. For the models developed by Barbieri et al.^16^ and Agmon et al.^17^, we also implemented changes to extract predicted probabilities of ICU readmission. Finally, by excluding patients who died during hospitalisation or within 30 days after discharge, we implicitly modified the original prediction tasks defined by Lin et al.^15^ and Agmon et al.^17^, which also incorporated mortality within 30 days of discharge or transfer to general wards as part of the outcome definition.

The modified codebases are available at https://github.com/ekoumantakis/ebma-icu-readmission.

#### Machine Learning models

Starting with our^5^ and others’^3,4^ systematic reviews, we again adapted six traditional machine learning models to ensure that all models were trained and evaluated on the same patient cohort. We selected four models from Lin et al.^15^ (*i*.*e*., a logistic regression with L1 regularization, a Naive Bayes, a Random Forest, and a Support Vector Machine), one model from Barbieri et al.^16^ (*i*.*e*., a logistic regression), and a model from Pakbin et al.^22^ (*i*.*e*., an XGBoost model). As for the deep learning models, these models were trained and evaluated on the same subset of patients (*i*.*e*., the training and test set). Of note, all models apart from the logistic regression from Barbieri et al.^16^ had their prediction task implicitly modified.

#### Statistical models

To include clinically validated prediction models, we also considered the Stability and Workload Index for Transfer (SWIFT)^23^ and Acute Physiologic Assessment and Chronic Health Evaluation (APACHE) II^24^ scores. SWIFT was specifically developed for the ICU readmission prediction task. APACHE was originally developed to predict death risk at ICU admission and subsequently adapted for the readmission task^25^. As scores evaluated before discharge are more predictive, we first extracted the most recent data recorded within the 72 (SWIFT) or 24 (APACHE II) or to discharge, and then assigned the corresponding points according to the original scoring systems. We considered missing values as not contributing to the total score (0 points). For the SWIFT score, we dichotomised the readmission risk into low (<15 points) and high (≥15 points), following the original study^23^. To derive predicted probabilities, we calculated the 30-day ICU readmission rates within the high- and low-risk groups in the training set, and used these as group-level probabilities, assigning them to patients in the calibration and test sets according to their risk category. As no validated threshold is available for APACHE II scores predicting ICU readmission, we treated them as a continuous predictor in a logistic regression model trained on the training set, and used it to estimate readmission probabilities in the calibration and test sets.

#### Model diversity

We generated predicted probabilities of ICU readmission for patients in the calibration set and computed pairwise Pearson correlation coefficients between these model outputs as a measure of prediction diversity.

### Ensemble Bayesian Model Averaging

We combined the predictions of the 25 individual models into a single estimate using an Ensemble Bayesian Model Averaging (EBMA)-based framework.

EBMA accounts for uncertainty about which candidate model best approximates the data generating process by assigning posterior weights to each model and combining their predictive distributions into a weighted ensemble prediction. The ensemble predictive distribution was represented as a finite mixture of the predictive distributions from the K candidate models:

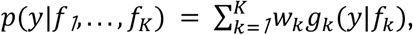

where *y* denotes the ICU readmission outcome; *f*_*k*_ is the prediction form model *k*, and *g*_*k*_(*y*|*f*_*k*_) is the predictive distribution under the model *k*. The quantity w_*k*_ is the posterior weight assigned to model *k*, with *w*_*k*_ ≥ *0* and 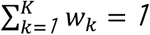.

For the binary outcome *y*_*i*_ ∈ {*0,1*}, each model *k* provided a predictive probability *p*_*ik*_ for observation *i* with Bernoulli likelihood 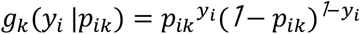.

The final EBMA-predicted probability for observation *i* was computed as a posterior mean over retained MCMC draws

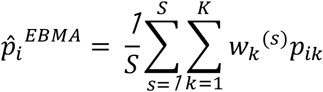

where *S* is the number of retained post-burn-in draws and 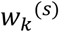 is the posterior draw of the weight assigned to model *k* at iteration *s*.

During calibration, performed exclusively on the held-out calibration set, EBMA weights were estimated via Gibbs sampling. The sampler was run for 40,000 iterations with the first 20,000 discarded as burn-in and every 20th subsequent draw retained. Model weights were initialized uniformly, with *W* = *1*/*k*. At each iteration, a latent allocation indicator *z*_*i*_ ∈ {*1, … k*}, representing the ensemble component associated with observation *i* in the mixture model representation, was sampled from a multinomial distribution.

After sampling, the allocation indicators, the weight vector *W* = (*w*_*1*_, *…, w*_*k*_) was updated by drawing from its full Dirichlet full conditional distribution, obtained by combining, a symmetric Dirichlet prior with the allocation counts across models. We summarized the posterior distribution of the ensemble weights using the retained post-burn-in, thinned MCMC draws. For each model we reported the posterior mean of *w*_*k*_ and the corresponding 95% CrI, calculated as the 2.5th and 97.5th percentiles of the posterior distribution.

We used the EBMA implemented in the *EBMAforecast* R package (version 1.0.32). As it cannot handle exact probability values for 0 or 1, these values were replaced with 1×10^−6^ and 1-1×10^−6^, respectively.

### Model evaluation

For each model, performance was evaluated on the test set, which was not used at any stage of model development or validation. Calibration was assessed using the Brier Score, which measures the mean squared difference between predicted probabilities and observed outcomes (ICU readmission *vs* no readmission). Discrimination was quantified using the area under the receiver operating characteristic curve (AUROC). As the training set differed from those used in the original studies, we assessed performance stability as the relative difference between the AUROC obtained in our analysis and that reported in the original studies, expressed as a proportion of the original AUROC.

### Computational Environment

EBMA calibration and evaluation ran on a laptop with an Intel i7-10875H 8-core processor and 16GB RAM, using R version 4.5.3. Figures were generated with the following R packages: *ggplot2* (v4.0.2), *pheatmap* (v1.0.13), and *ggridges* (v0.5.7).

## Declarations

### Data availability

MIMIC-III is publicly available upon a motivated request to Physionet (https://physionet.org/content/mimiciii/1.4/). Due to the sensitive nature of the data, which contain potentially identifiable health information, prediction probabilities for patients in the calibration and test sets will be made available from the corresponding author upon reasonable request to *bona fide* researchers, subject to appropriate ethical approvals and data use agreements.

### Code availability

The code supporting this study is publicly available at: https://github.com/ekoumantakis/ebma-icu-readmission.

### Contributions

EK, AV, and PB conceptualised and designed the study. EK curated the MIMIC-III dataset. EK adapted and trained the models selected for EBMA integration. EK and AV assessed model diversity, calibrated the EBMA-based framework, and performed model evaluation. EK generated Figure 1, and AV generated the remaining figures. EK, AV, and PB interpreted the results. EK and AV wrote the manuscript with contributions from PB, KR, and IR. All authors critically reviewed, edited, and approved the final version.

## Abbreviations

ANOVA: Analysis Of Variance
APACHE II: Acute Physiologic Assessment and Chronic Health Evaluation II
AUROC: Area Under the Receiver Operating Characteristic Curve
CNN: Convolutional Neural Network
CrI: Credible Interval
EBMA: Ensemble Bayesian Model Averaging
ICU: Intensive Care Unit
IQR: interquartile range
LSTM: Long Short-Term Memory
MCE: Medical Concept Embeddings
MIMIC-III: Medical Information Mart for Intensive Care III
ODE: Ordinary Differential Equation
RNN: Recurrent Neural Network
SD: standard deviation
SWIFT: Stability and Workload Index for Transfer
TeDi-BERT: Temporal Distribution matching training method, applied to Bidirectional Encoder Representations from Transformers
XGBoost: eXtreme Gradient Boosting

