## Supplementary Material for "Improving machine learning and deep learning models for 30-day ICU readmission prediction using Ensemble Bayesian Model Averaging"

### Supplementary Figures

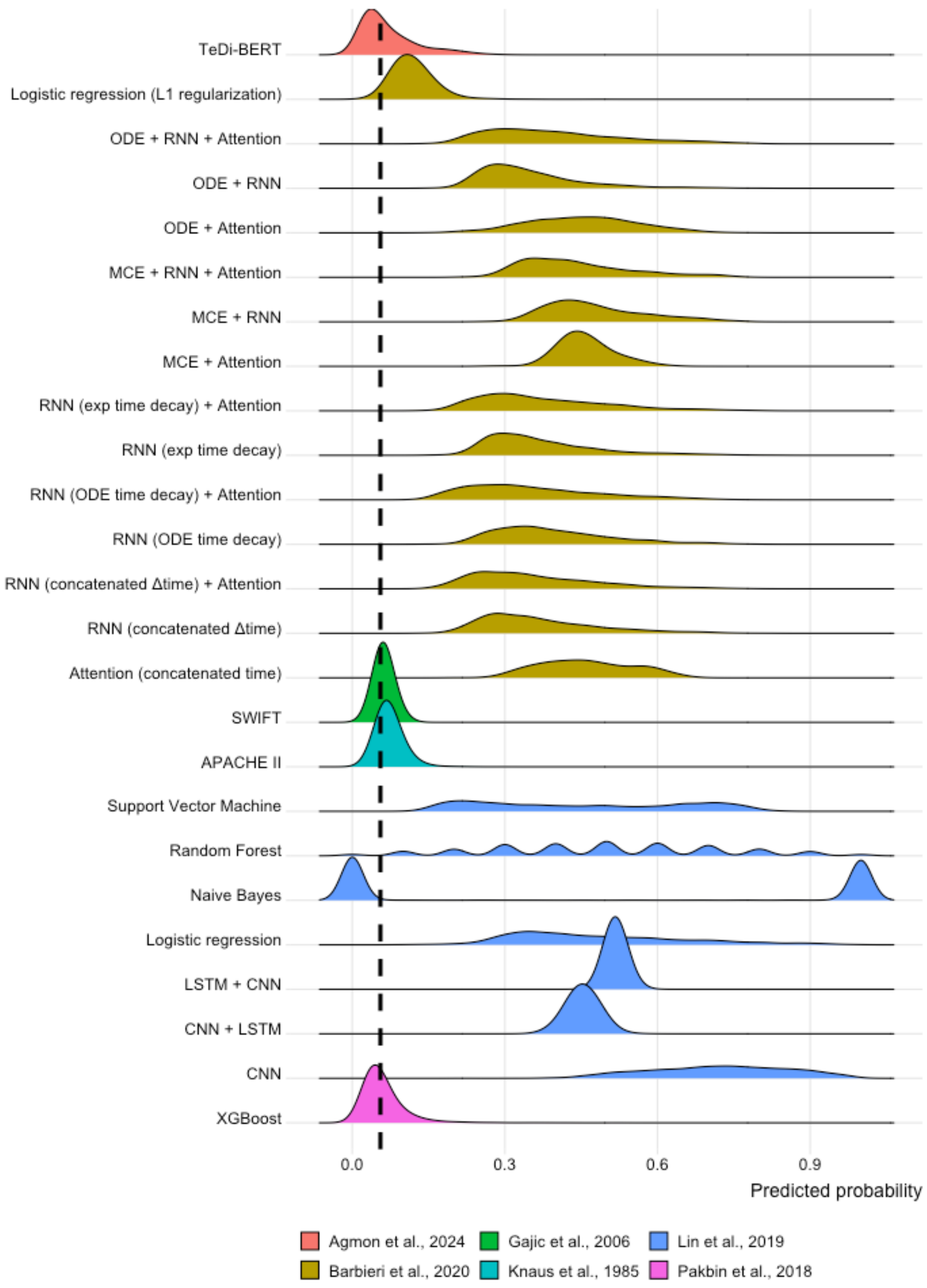


**Supplementary Figure 1. Ridge plot of predicted probability distributions (individual models).** Each ridge represents the density of predicted probabilities of the 25 individual models for patients in the test set. The vertical dotted line shows the expected ICU readmission prevalence (*i.e.*, 5.5%). Colours indicate original studies. *Abbreviations: CNN, Convolutional Neural Network; APACHE II, Acute Physiologic Assessment and Chronic Health Evaluation II; CNN, Convolutional Neural Network; LSTM, Long Short-Term Memory; MCE, Medical Concept Embeddings; ODE, Ordinary Differential Equation; RNN, Recurrent Neural Network; SWIFT, Stability and Workload Index for Transfer; TeDi-BERT, Temporal Distribution matching training method, applied to Bidirectional Encoder Representations from Transformers; XGBoost, eXtreme Gradient Boosting.*

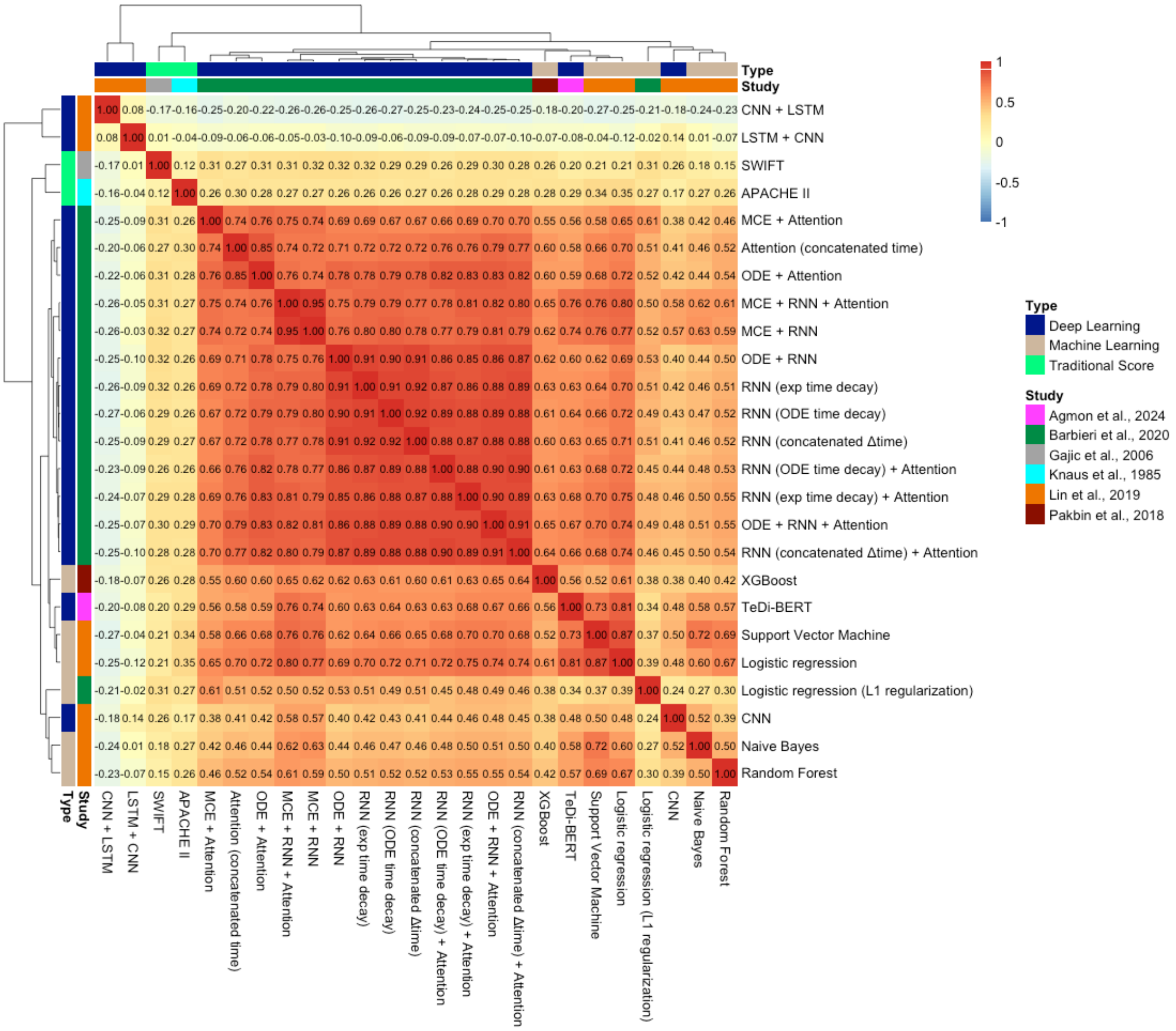


**Supplementary Figure 2. Model diversity.** The heatmap shows the pairwise Pearson correlation coefficients between predicted probabilities for the 25 individual models. Values close to -1 (blue) indicate perfect negative correlation, while values close to 1 (red) perfect positive correlation. Near-zero values (white) indicate no correlation. Rows and columns are clustered using Euclidean distance, and the resulting dendrogram is also shown. Colour bars indicate original studies and model type (*i.e.*, deep learning, machine learning, and traditional scores)

*Abbreviations: APACHE II, Acute Physiologic Assessment and Chronic Health Evaluation II; CNN, Convolutional Neural Network; LSTM, Long Short-Term Memory; MCE, Medical Concept Embeddings; ODE, Ordinary Differential Equation; RNN, Recurrent Neural Network; SWIFT, Stability and Workload Index for Transfer; TeDi-BERT, Temporal Distribution matching training method, applied to Bidirectional Encoder Representations from Transformers; XGBoost, eXtreme Gradient Boosting.*

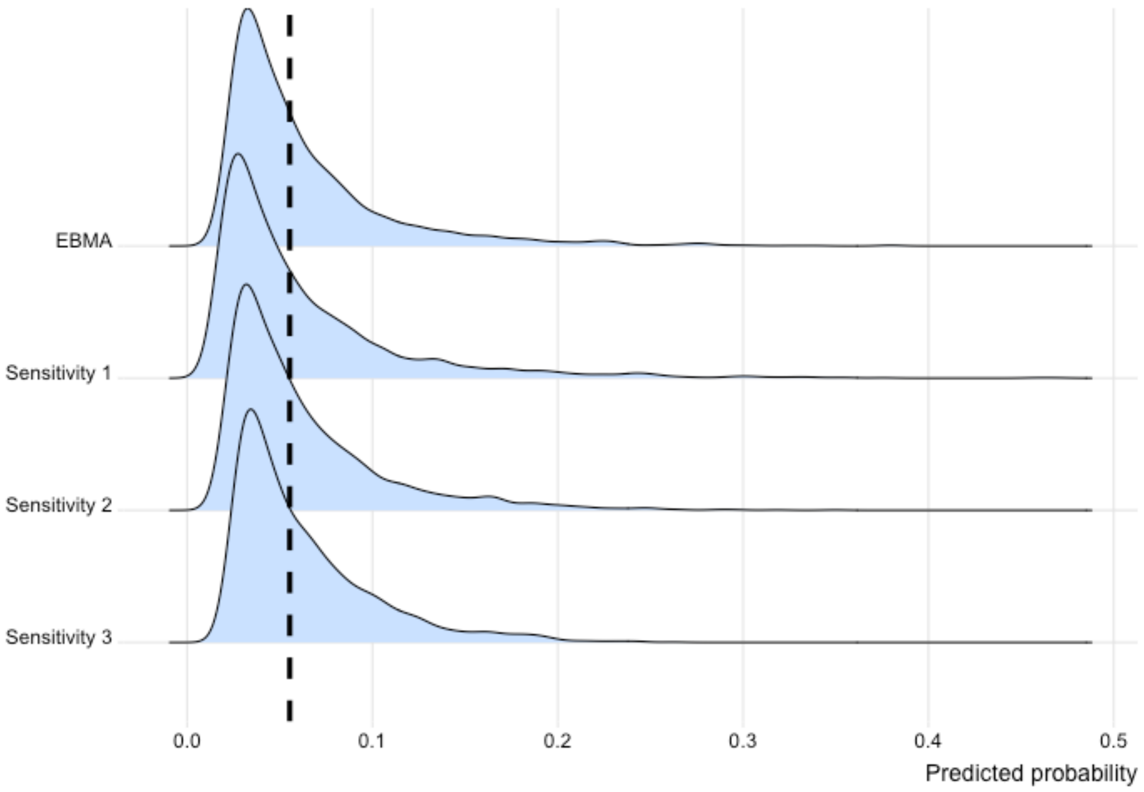


**Supplementary Figure 3. Ridge plot of predicted probability distributions (EBMA-based framework).** Each ridge represents the density of predicted probabilities of the EBMA-based framework for patients in the test set, including during sensitivity analyses. EBMA indicates the complete EBMA-based framework including all 25 models. Sensitivity 1 excluded models whose weight in the original framework was lower than an uninformative prior (<1/25=0.04). Sensitivity 2 excluded models whose pairwise Pearson correlation coefficient was larger than 0.8. Sensitivity 3 further excluded the strongest individual contributor from those included in Sensitivity 2. The vertical dotted line shows the expected ICU readmission prevalence (*i.e.*, 5.5%).
*Abbreviations: EBMA, Ensemble Bayesian Model Averaging.*

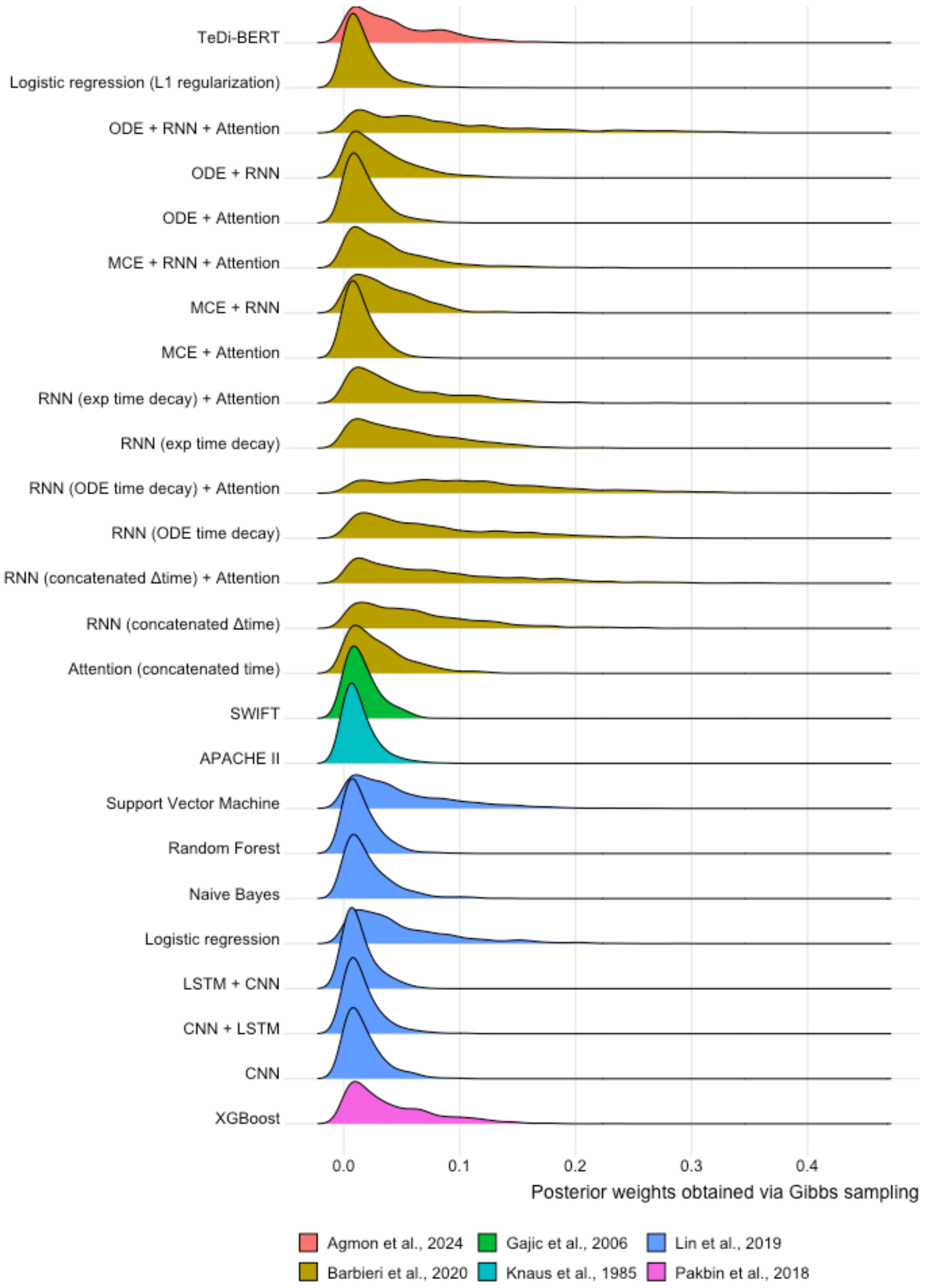


**Supplementary Figure 4. Posterior distribution of EBMA weights.** Each ridge represents the posterior weight distribution of the 25 individual models included in the EBMA-based framework. Colours indicate original studies.

*Abbreviations: APACHE II, Acute Physiologic Assessment and Chronic Health Evaluation II; CNN, Convolutional Neural Network; LSTM, Long Short-Term Memory; MCE, Medical Concept Embeddings; ODE, Ordinary Differential Equation; RNN, Recurrent Neural Network; SWIFT, Stability and Workload Index for Transfer; TeDi-BERT, Temporal Distribution matching training method, applied to Bidirectional Encoder Representations from Transformers; XGBoost, eXtreme Gradient Boosting.*

### Supplementary Tables

**Supplementary Table 1. Performance of the 25 individual models.** The table reports the performance of the individual models in the test set, evaluated using the Brier score (calibration) and the area under the receiver operating characteristic curve (AUROC; discrimination). The AUROC values reported in the original studies (AUROC*) are also provided, together with the relative difference between the originally reported and observed AUROC values in this study, expressed as a proportion of the original AUROC (Difference).

| Study | Model architecture | Brier score | AUROC | AUROC* | Difference |
|---|---|---:|---:|---:|---:|
| (Agmon et al., 2024) | TeDi-BERT | 0.051 | 0.684 | 0.650 | 5.2% |
| (Barbieri et al., 2020) | RNN (ODE time decay) | 0.171 | 0.712 | 0.741 | -3.9% |
| | RNN (concatenated Δtime) + Attention | 0.148 | 0.716 | 0.741 | -3.4% |
| | RNN (ODE time decay) + Attention | 0.148 | 0.714 | 0.743 | -3.9% |
| | ODE + RNN + Attention | 0.176 | 0.706 | 0.739 | -4.5% |
| | RNN (concatenated Δtime) | 0.153 | 0.714 | 0.739 | -3.4% |
| | RNN (exp time decay) + Attention | 0.160 | 0.700 | 0.748 | -6.4% |
| | MCE + RNN | 0.232 | 0.709 | 0.727 | -2.5% |

| | | | | | |
|---|---|---|---|---|---|
| | MCE + RNN + Attention | 0.205 | 0.699 | 0.736 | -5.0% |
| | RNN (exp time decay) | 0.155 | 0.702 | 0.735 | -4.5% |
| | Attention (concatenated time) | 0.220 | 0.672 | 0.711 | -5.5% |
| | ODE + RNN | 0.140 | 0.701 | 0.739 | -5.1% |
| | ODE + Attention | 0.215 | 0.671 | 0.717 | -6.4% |
| | MCE + Attention | 0.215 | 0.667 | 0.689 | -3.2% |
| | Logistic regression | 0.056 | 0.640 | 0.659 | –2.9% |
| (Lin et al., 2019) | CNN | 0.507 | 0.621 | 0.784 | -20.8% |
| | CNN + LSTM | 0.212 | 0.442 | 0.787 | -43.8% |
| | LSTM + CNN | 0.266 | 0.500 | 0.791 | -36.8% |
| | Logistic regression (L1 regularization) | 0.254 | 0.697 | 0.777 | -10.3% |
| | Naive Bayes | 0.455 | 0.636 | 0.706 | -9.9% |

| | | | | | |
|---|---|---|---|---|---|
| | Support Vector Machine | 0.236 | 0.712 | 0.779 | -8.6% |
| | Random Forest | 0.287 | 0.649 | 0.712 | -8.8% |
| (Pakbin et al., 2018) | XGBoost | 0.052 | 0.675 | 0.750 | -10.0% |
| (Gajic et al., 2008) | SWIFT score | 0.052 | 0.530 | 0.750 | -29.3% |
| (Knaus et al., 1985) | APACHE II score | 0.052 | 0.653 | 0.606[a] | 7.8% |

[a]As reported in [Lee *et al.*, 2015; doi: 10.1177/0310057X1504300206], which adapted the APACHE II score for the ICU-readmission task

*Abbreviations: APACHE II, Acute Physiologic Assessment and Chronic Health Evaluation II; CNN, Convolutional Neural Network; LSTM, Long Short-Term Memory; MCE, Medical Concept Embeddings; ODE, Ordinary Differential Equation; RNN, Recurrent Neural Network; SWIFT, Stability and Workload Index for Transfer score; TeDi-BERT, Temporal Distribution matching training method, applied to Bidirectional Encoder Representations from Transformers; XGBoost, eXtreme Gradient Boosting.*

**Supplementary Table 2. Uncertainty of EBMA posterior weights.** The table reports, for each individual model, the mean posterior weight (w) and the corresponding 95% credible interval (95% CrI; defined as the 2.5th and 97.5th percentiles of the posterior weight distribution). EBMA indicates the complete EBMA-based framework. Sensitivity 1 excluded models whose weights in the original framework were lower than the uninformative prior (<1/25=0.04). Sensitivity 2 excluded models with pairwise Pearson correlation coefficient greater than 0.8. Sensitivity 3 further excluded the strongest individual contributor among the models retained in Sensitivity 2.

| **Study** | **Model architecture** | **EBMA** | | **Sensitivity 1** | | **Sensitivity 2** | | **Sensitivity 3** | |
|---|---|---|---|---|---|---|---|---|---|
| | | **w** | **95% CrI** | **w** | **95% CrI** | **w** | **95% CrI** | **w** | **95%CrI** |
| (Agmon et al., 2024) | TeDi-BERT | 0.044 | 0.001–0.137 | 0.061 | 0.001-0.177 | 0.069 | 0.002-0.211 | 0.110 | 0.005-0.295 |
| (Barbieri et al., 2020) | RNN (ODE time decay) | 0.075 | 0.004–0.230 | 0.149 | 0.004-0.377 | - | - | - | - |
| | RNN (concatenated Δtime) + Attention | 0.078 | 0.003–0.256 | 0.138 | 0.008-0.373 | - | - | - | - |
| | RNN (ODE time decay) + Attention | 0.113 | 0.007–0.307 | 0.154 | 0.005-0.376 | 0.453 | 0.277-0.629 | - | - |
| | ODE + RNN + Attention | 0.086 | 0.003–0.293 | 0.108 | 0.005-0.316 | - | - | - | - |
| | RNN (concatenated Δtime) | 0.063 | 0.003–0.204 | 0.064 | 0.001-0.201 | - | - | - | - |

| | | | | | | | | | |
|---|---|---|---|---|---|---|---|---|---|
| | RNN (exp time decay) + Attention | 0.050 | 0.001–0.159 | 0.053 | 0.002-0.160 | - | - | - | - |
| | MCE + RNN | 0.037 | 0.001–0.120 | - | - | - | - | - | - |
| | MCE + RNN + Attention | 0.039 | 0.001–0.154 | - | - | 0.089 | 0.002-0.302 | 0.268 | 0.055-0.502 |
| | RNN (exp time decay) | 0.053 | 0.001–0.154 | 0.079 | 0.003-0.229 | - | - | - | - |
| | Attention (concatenated time) | 0.030 | 0.001–0.103 | - | - | 0.043 | 0.001-0.183 | 0.132 | 0.004-0.334 |
| | ODE + RNN | 0.031 | 0.001–0.104 | - | - | - | - | - | - |
| | ODE + Attention | 0.017 | 0.000–0.063 | - | - | - | - | - | - |
| | MCE + Attention | 0.014 | 0.000–0.047 | - | - | 0.017 | 0.001-0.060 | 0.026 | 0.001-0.097 |
| | Logistic regression | 0.017 | 0.001–0.063 | - | - | 0.021 | 0.001-0.071 | 0.023 | 0.001-0.080 |
| (Lin et al., 2019) | CNN | 0.017 | 0.000–0.060 | - | - | 0.022 | 0.001-0.076 | 0.027 | 0.001-0.093 |
| | CNN + LSTM | 0.016 | 0.000–0.060 | - | - | 0.022 | 0.001-0.078 | 0.020 | 0.001-0.071 |

| | | | | | | | | | |
|---|---|---|---|---|---|---|---|---|---|
| | LSTM + CNN | 0.013 | 0.000–0.049 | - | - | 0.014 | 0.000-0.045 | 0.018 | 0.001-0.063 |
| | Logistic regression (L1 regularization) | 0.049 | 0.002–0.167 | 0.104 | 0.006-0.277 | - | - | - | - |
| | Naive Bayes | 0.021 | 0.001–0.082 | - | - | 0.029 | 0.000-0.103 | 0.036 | 0.001-0.117 |
| | Support Vector Machine | 0.049 | 0.001–0.163 | 0.090 | 0.003-0.257 | 0.089 | 0.005-0.257 | 0.155 | 0.009-0.345 |
| | Random Forest | 0.015 | 0.000–0.052 | - | - | 0.022 | 0.000-0.088 | 0.021 | 0.001-0.072 |
| (Pakbin et al., 2018) | XGBoost | 0.040 | 0.001–0.131 | - | - | 0.079 | 0.003-0.221 | 0.126 | 0.007-0.299 |
| (Gajic et al., 2008) | SWIFT score | 0.016 | 0.000–0.053 | - | - | 0.013 | 0.000-0.050 | 0.017 | 0.001-0.069 |
| (Knaus et al., 1985) | APACHE II score | 0.013 | 0.000–0.051 | - | - | 0.018 | 0.000-0.068 | 0.022 | 0.000-0.083 |

*Abbreviations: APACHE II, Acute Physiologic Assessment and Chronic Health Evaluation II; CNN, Convolutional Neural Network; LSTM, Long Short-Term Memory; MCE, Medical Concept Embeddings; ODE, Ordinary Differential Equation; RNN, Recurrent Neural Network; SWIFT, Stability and Workload Index for Transfer score; TeDi-BERT, Temporal Distribution matching training method, applied to Bidirectional Encoder Representations from Transformers; XGBoost, eXtreme Gradient Boosting*
